# Association of heavy menstrual bleeding with cardiovascular diseases in US female hospitalizations

**DOI:** 10.1101/2023.09.11.23295385

**Authors:** Pallavi Dubey, Jasmin Abdeldayem, Vishwajeet Singh, Yousif Abdelrehman, Sanjana Mada, Sireesha Reddy, Alok Dwivedi

**Affiliations:** Department of Obstetrics and Gynecology, Paul L. Foster School of Medicine, Texas Tech University Health Sciences Center El Paso, El Paso, Texas, US; Paul L. Foster School of Medicine, Office of Research Resources, Biostatistics and Epidemiology Consulting Lab, Texas Tech University Health Sciences Center El Paso, El Paso, Texas, USA; Division of Biostatistics & Epidemiology, Department of Molecular and Translational Medicine, Paul L. Foster School of Medicine, Texas Tech University Health Sciences Center El Paso, El Paso, Texas, USA; Paul L. Foster School of Medicine, Texas Tech University Health Sciences Center El Paso, El Paso, Texas, US

**Keywords:** menorrhagia, heavy menstrual bleeding, cardiovascular disease, irregular menstruation, obesity, metabolic syndrome

## Abstract

**BACKGROUND:** Heavy menstrual bleeding (HMB) is a common type of menstrual disorder that causes social and mental distress leading to a variety of health problems. Although limited evidence highlights the links between HMB and metabolic abnormalities, the association between HMB and CVD outcomes is unknown in the presence or absence of comorbid irregular menstruation (IM).

**METHODS:** All female hospitalizations between the ages of 12 to 40 years were identified using the National Inpatient Sample database in 2017 and grouped into HMB and no menstrual disorders. Outcomes including major adverse cardiovascular events (MACE), coronary heart disease (CHD), stroke/cerebrovascular accident (CVA), heart failure (HF), arterial fibrillation (AF) or arrhythmia, myocardial infarction (MI), and diabetes (DM) were extracted using ICD-10 codes. Adjusted logistic regression models were conducted to summarize associations with an odds ratio (OR) along with a 95% confidence interval (CI).

**RESULTS:** In the entire cohort, 8,291 (0.69%) had an HMB diagnosis including 1623 (0.14%) with IM and 6668 (0.55%) without IM. In the fully adjusted analysis, HMB was significantly associated with most CVD outcomes, including MACE (OR=1.61; 95% CI: 1.38, 1.88, p<0.001), CHD (OR=1.88; 95% CI: 1.52, 2.31, p<0.001), Stroke/CVA (OR=1.51; 95% CI: 1.1, 2.07, p=0.011), HF (OR=1.60; 95% CI: 1.33, 1.92, p<0.001), AF/arrhythmia (OR=1.93; 95% CI: 1.16, 2.45, p=0.006), and DM (OR=1.45; 95% CI: 1.33, 1.58), p<0.001). HMB without IM was strongly associated with DM, HF, AF, and MACE outcomes while HMB with IM was strongly associated with CHD and AF outcomes.

**CONCLUSIONS:** HMB is independently and strongly associated with CVD events among US hospitalizations of young women. Our findings suggest promoting increased awareness, early detection, and optimum management of HMB to avoid adverse CVD outcomes.

**Clinical Perspective:** *What Is New?:*

- Although menstruation dysfunction is associated with cardiovascular disease (CVD), the relationship between heavy menstrual bleeding (HMB) and CVD outcomes is not evaluated.
- In a US sample of 1,204,715 hospitalized women, HMB was independently and strongly associated with major adverse cardiovascular events and coronary heart disease.
- HMB without irregular menstruation was markedly associated with CVD outcomes than HMB with irregular menstruation.

*What Are the Clinical Implications?:* Our findings support that HMB as an early vital sign for CVD outcomes among young women. A routine investigation and screening of menstrual disorders, especially HMB, and its effective management is useful for the prevention of CVD risk in women.

## INTRODUCTION

Cardiovascular disease (CVD) is the leading cause of mortality with a global burden of approximately 19 million deaths in 2020, accounting for about 32% of all global deaths.^1^ Considering sex differences and the increasing prevalence of metabolic syndrome and CVD, particularly in women with menstrual disorders, it becomes critical to identify modifiable risk factors for CVD risk prevention in women.^2, 3^ Menstrual disorders refer to a wide range of conditions that impact a woman’s menstrual cycle, affecting several factors such as regularity, duration, severity, or other associated symptoms yielding severe complications and adverse health outcomes.^4–6^ Heavy menstrual bleeding (HMB), a type of menstrual disorder, involves excessive menstrual blood loss of greater than 80 mL per menstrual period or clinically excessive bleeding that affects physical, emotional, and social distress, and material quality of life of affected females.^7^ HMB is one of the prevalent menstrual disorders of reproductive-aged women affecting 9% to 52% of women depending on objective and subjective assessments of HMB.^8^ HMB is also referred to as menorrhagia and is sometimes interchanged with abnormal uterine bleeding as it often occurs due to uterine/endometrial, hormonal, or coagulation-related conditions.^9^ It is typically associated with anemia, fatigue, headache, and pain.^10^ Its association with iron deficiency anemia can impair oxygen transport and interfere with cardiac function.^11, 12^ In addition, hormonal-related changes and other associated morbidities of HMB may adversely affect cardiometabolic health leading to CVD outcomes.^10, 13^

To address sex differences in CVD in women, irregular menstruation (IM), premature menopause, and long menstrual cycles have been studied as risk factors for CVD outcomes.^2^ Since IM is considered the prominent feature of polycystic ovary syndrome (PCOS)^14^, a condition strongly associated with cardiometabolic health^15^, several studies examined the association of IM with CVD outcomes and mortality.^5, 16, 17^ In small-sized studies, HMB was also found to be associated with secondary PCOS, insulin resistance, obesity, and inflammation indicating that HMB can be associated with CVD.^18–20^ Approximately one-third of gynecological visits in the US are due to HMB and represent a heavy economic burden on patients for treatment costs and productivity loss.^8, 21^ Although HMB is a prevalent condition associated with hormonal changes, obesity, arrhythmia, hypertension, and anemia that may increase the risk of heart disease^10, 20, 22, 23^, a direct association between HMB and CVD outcomes including coronary heart disease (CHD), heart failure (HF), stroke, atrial fibrillation (AF)/arrhythmia, myocardial infarction (MI), and diabetes (DM) has not been evaluated among young women. We sought to determine the association of HMB with or without IM with multiple CVD outcomes using a nationally representative sample of female hospitalizations in the US.

## METHODS

### Study Population

We conducted a retrospective cross-sectional using the National Inpatient Sample (NIS) database from 2017. The NIS is a publicly available all-payer inpatient healthcare database designed to produce U.S. regional and national estimates of inpatient outcomes, resources, and costs. The NIS data collection and procedures followed Helsinki’s declaration of ethical standards. The NIS, 2017 includes de-identified datasets from the International Classification of Diseases, 10th Revision (ICD-10) codes from participating hospitals and hence, does not require participant consent or approval. In our study, we included all women with excessive and frequent menstruation with regular cycles (ICD: N920) or without regular cycles (ICD: N921) of ages between 12-40 years. We excluded hospitalized women with hematocolpos, amenorrhea, excessive menstruation at puberty, ovulation bleeding, irregular menstruation, and dysmenorrhea.

### Outcomes

The study outcomes were major adverse cardiovascular events (MACE), CHD, stroke/cardiovascular accident (CVA), HF, AF or arrhythmia, MI, and DM. Due to limited CVD mortality and no coronary revascularization, MACE was defined as the presence of any events related to stroke, MI, and HF.^15^ These outcomes were defined using the ICD-10 and related ICD codes were provided in another publication.^15^ The primary exposure variable was HMB with or without IM.

### Covariates

The covariates considered in this study were age (12-20, 21-30, and 31-40 years), race/ethnicity (white, black, Hispanic, and other/missing), household (HH) income quartile (1^st^, 2^nd^, 3^rd^, 4^th^, and missing), primary payer (Medicare, Medicaid, private insurance, self-pay, no charge, and others/missing), smoking status (no and yes), alcohol (no and yes), obesity (no and yes), and metabolic syndrome (no and yes). Metabolic syndrome (MS) was defined as the presence of at least three conditions among fasting glucose, low high-density lipoprotein, high blood pressure, high triglyceride, and obesity.^15^ For validation analysis, we also collected data on PCOS, congenital heart disease, and anticoagulation use using the appropriate ICD codes.

### Statistical Analysis

All the statistical analyses were carried out by applying the appropriate sampling weight in accordance with the Healthcare Cost and Utilization Project (HCUP)-NIS protocol.^15^ Data were summarized with appropriate descriptive statistics including mean with standard deviation or frequencies with percentages. The association between HBM and each CVD outcome was evaluated using survey-weighted logistic regression analyses. We explored all possible interactions with HMB prior to finalizing the variables that need to be adjusted in the adjusted analysis. Accordingly, the considered covariates i.e. race/ethnicity, household income quartile, primary payer, smoking status, and alcohol were adjusted in the analyses as per the study objective.^24^ The results of logistic regression were summarized with odds ratio (OR) with 95% confidence interval (CI). The associations were also validated using modified Poisson regression models to estimate the relative risk (RR) with 95%CI. We performed various sensitivity analyses to confirm our study findings. Multiple logistic regression models were developed after additionally adjusting for obesity, metabolic syndrome, and diabetes separately from the primary adjusted model. Further, the results were validated after excluding the women with PCOS hospitalizations, congenital heart disease, and anticoagulation use. Although age had an interaction effect with HMB, we also summarized the adjusted association of HMB with CVD outcomes after additionally adjusting with age along with other covariates using survey-weighted logistic regression analysis. We further evaluated the association between HMB with or without IM and each CVD outcome using survey-weighted logistic and Poisson regression analyses. STATA 17.0 version was used for the data management and statistical analyses. A p-value less than 5% was considered a statistically significant result. We followed statistical analysis and methods in biomedical research (SAMBR) checklists and crafted statistical analysis as per the guidance resources.^24^

## RESULTS

A total of 1,204,715 hospitalized women records were found to be eligible for statistical analysis. The mean (SD) age of the female hospitalizations was 28.45 (6.51) years, with a majority in the age group of 21-30 (46.19%) and 31-40 (41.23%). Half of the hospitalizations were non-Hispanic white with an equal distribution of non-Hispanic black (17.26%) and Hispanic (17.92%). One-third of the hospitalized women (30.23%) had household income in the first quartile and primary payers were mainly private (45.65%) or Medicaid (43.02%). A limited proportion of hospitalized women (0.57%) were smokers with 2.2% alcoholic drinkers, 12.8% obese, and 1.1% with metabolic syndrome (Table 1). In the total cohort, 0.69% of hospitalizations (n=8291) had a diagnosis of HMB which includes 1623 (0.14%) hospitalizations with IM and 6668 (0.55%) without IM (Table 1).

**Table 1.** Sample characteristics of HMB among hospitalized women of age 12-40 years, NIS-2017.

| **Characteristics** | **Total** | **No HMB** | **HMB** | **p-value** |
| --- | --- | --- | --- | --- |
| **Total** | **N=1204715** | **N=1196424** | **N=8291** |  |
| **Age (years)** |  |  |  | <0.001 |
| 12-20 | 151515 (12.58%) | 150655 (12.59%) | 860 (10.37%) |  |
| 21-30 | 556478 (46.19%) | 554714 (46.36%) | 1764 (21.28%) |  |
| 31-40 | 496722 (41.23%) | 491055 (41.04%) | 5667 (68.35%) |  |
| Mean (SD) | 28.45 (6.51) | 28.43 (6.50) | 32.03 (7.19) |  |
| **Race/Ethnicity** |  |  |  | <0.001 |
| White | 610872 (50.71%) | 607970 (50.82%) | 2902 (35.00%) |  |
| Black | 207962 (17.26%) | 204928 (17.13%) | 3034 (36.59%) |  |
| Hispanic | 215924 (17.92%) | 214462 (17.93%) | 1462 (17.63%) |  |
| Others/Missing | 169957 (14.11%) | 169064 (14.13%) | 893 (10.77%) |  |
| **HH income quartile** |  |  |  | <0.001 |
| 1st | 364132 (30.23%) | 361082 (30.18%) | 3050 (36.79%) |  |
| 2nd | 310548 (25.78%) | 308366 (25.77%) | 2182 (26.32%) |  |
| 3rd | 280497 (23.28%) | 278718 (23.30%) | 1779 (21.46%) |  |
| 4th | 235121 (19.52%) | 233948 (19.55%) | 1173 (14.15%) |  |
| Missing | 14417 (1.20%) | 14310 (1.20%) | 107 (1.29%) |  |
| **Primary payer** |  |  |  | <0.001 |
| Medicare | 41678 (3.46%) | 41312 (3.45%) | 366 (4.41%) |  |
| Medicaid | 518302 (43.02%) | 515339 (43.07%) | 2963 (35.74%) |  |
| Private insurance | 549941 (45.65%) | 546011 (45.64%) | 3930 (47.40%) |  |
| Self-Pay | 53708 (4.46%) | 52998 (4.43%) | 710 (8.56%) |  |
| No charge | 3222 (0.27%) | 3154 (0.26%) | 68 (0.82%) |  |
| Other | 37864 (3.14%) | 37610 (3.14%) | 254 (3.06%) |  |
| **Smoking status** |  |  |  | <0.001 |
| No | 1197802 (99.43%) | 1189600 (99.43%) | 8202 (98.93%) |  |
| Yes | 6913 (0.57%) | 6824 (0.57%) | 89 (1.07%) |  |
| **Alcohol** |  |  |  | 0.58 |
| No | 1178369 (97.81%) | 1170252 (97.81%) | 8117 (97.90%) |  |
| Yes | 26346 (2.19%) | 26172 (2.19%) | 174 (2.10%) |  |
| **Obesity** |  |  |  | <0.001 |
| No | 1050043 (87.16%) | 1043629 (87.23%) | 6414 (77.36%) |  |
| Yes | 154672 (12.84%) | 152795 (12.77%) | 1877 (22.64%) |  |
| **MS** |  |  |  | <0.001 |
| No | 1191459 (98.90%) | 1183412 (98.91%) | 8047 (97.06%) |  |
| Yes | 13256 (1.10%) | 13012 (1.09%) | 244 (2.94%) |  |

Abbreviations: NIS, National inpatient sample; SD, Standard deviation; HH, Household; MS: Metabolic syndrome.

All the background characteristics were found to be different between HMB and no HMB hospitalizations except for alcohol status. HMB was strongly associated with age and black race. The proportion of obesity (22.6% vs. 12.8%, p<0.001) and MS (2.94% vs. 1.09%$, p<0.001) was significantly higher in HMB hospitalizations than in non-HMB hospitalizations (Table 1). The proportion of MACE was found highest (1.35%, n=16289) after diabetes (5.06%, n=60962) in our selected cohort, and the lowest proportion was observed for MI with 0.17% (n=2063). All the CVD outcomes were significantly higher in proportions in the HMB group compared to the non-HMB group (Table S1).

### Unadjusted and adjusted associations of HMB diagnosis with CVD outcomes

In the unadjusted analyses, the odds of MACE, CHD, HF, AF/arrhythmia, and MI were 2-fold higher in HMB hospitalizations compared to non-HMB hospitalizations. A 70% or higher odds of stroke and DM were observed in HMB hospitalizations compared to non-HMB hospitalizations. (Table 2). There was a strong interaction effect of age and HMB on CVD outcomes reflecting an age-dependent association between HBM and CVD outcomes. All the CVD outcomes except MI were strongly associated with HMB in the adjusted analysis accounting for age interaction with HMB (Table S2). After adjusting for race/ethnicity, household income quartile, primary payer, alcohol intake, and smoking status, HMB was found to be associated with MACE (OR=1.61; 95% CI: 1.38, 1.88, p<0.001), CHD (OR=1.88; 95% CI: 1.52, 2.31, p<0.001), Stroke/CVA (OR=1.51; 95% CI: 1.1, 2.07, p=0.011), HF (OR=1.60; 95% CI: 1.33, 1.92, p<0.001), AF/arrhythmia (OR=1.93; 95% CI: 1.16, 2.45, p=0.006), and DM (OR=1.45; 95% CI: 1.33, 1.58), p<0.001) (Table 2). The relative risk models also yielded the same pattern of effect sizes in the adjusted analyses (Figure 1). HMB without IM was only associated with MACE (OR=1.69; 95%CI: 1.43, 2.00, p<0.001), stroke/CVA (OR=1.53; 95%CI: 1.08, 2.17), HF (OR=1.68; 95%CI: 1.38, 2.05), and MI (OR=1.80; 95%CI: 1.20, 2.69). However, HMB with IM was associated with CHD (OR=2.10; 95%CI: 1.39, 3.18) to a greater extent than HMB without IM (OR=1.82; 95%CI: 1.43, 2.31) (Table 3).

**Figure 1.**
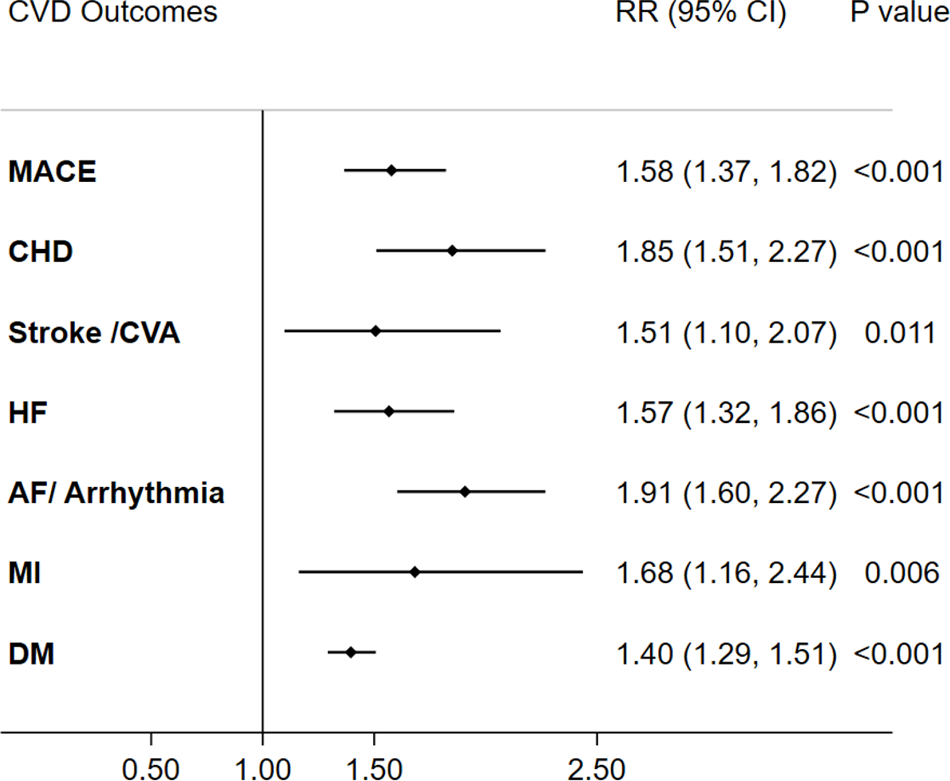
Adjusted relative risk for the association between HMB and each CVD outcome including diabetes among hospitalized women of ages between 12-40 years. RR, Relative risk; CI: Confidence interval; MACE, Major adverse cardiovascular event; CHD, coronary heart disease; CVA, Cerebrovascular accident; HF, Heart failure; AF, Arterial fibrillation; MI, Myocardial infarction; DM, Diabetes.

**Table 2.** Unadjusted and adjusted associations of HMB with CVD outcomes including diabetes among hospitalized women of ages between 12-40 years, NIS-2017.

|  | Unadjusted model |  | Adjusted model* |  |
| --- | --- | --- | --- | --- |
|  | OR (95% CI) | p-value | OR (95% CI) | p-value |
| **MACE** |  |  |  |  |
| No HMB (reference) | 1 |  | 1 |  |
| HMB | 2.03 (1.75, 2.35) | <0.001 | 1.61 (1.38, 1.88) | <0.001 |
| **CHD** |  |  |  |  |
| No HMB (reference) | 1 |  | 1 |  |
| HMB | 2.22 (1.82, 2.72) | <0.001 | 1.88 (1.52, 2.31) | <0.001 |
| **Stroke /CVA** |  |  |  |  |
| No HMB (reference) | 1 |  | 1 |  |
| HMB | 1.75 (1.28, 2.40) | <0.001 | 1.51 (1.10, 2.07) | 0.011 |
| **HF** |  |  |  |  |
| No HMB (reference) | 1 |  | 1 |  |
| HMB | 2.06 (1.73, 2.45) | <0.001 | 1.60 (1.33, 1.92) | <0.001 |
| **AF/ Arrhythmia** |  |  |  |  |
| No HMB (reference) | 1 |  | 1 |  |
| HMB | 2.07 (1.73, 2.46) | <0.001 | 1.93 (1.61, 2.3) | <0.001 |
| **MI** |  |  |  |  |
| No HMB (reference) | 1 |  | 1 |  |
| HMB | 1.99 (1.38, 2.87) | <0.001 | 1.69 (1.16, 2.45) | 0.006 |
| **DM** |  |  |  |  |
| No HMB (reference) | 1 |  | 1 |  |
| HMB | 1.69 (1.56, 1.84) | <0.001 | 1.45 (1.33, 1.58) | <0.001 |

Abbreviations: NIS, National inpatient sample; OR, Odds ratio; CI, Confidence interval; MACE, Major adverse cardiovascular event; CHD, coronary heart disease; CVA, Cerebrovascular accident; HF, Heart failure; AF, Arterial fibrillation; MI, Myocardial infarction; DM, Diabetes; HMB, Heavy menstrual bleeding.
MACE was defined as the composite of myocardial infarction, stroke, and heart failure.
\*Adjusted model included race/ethnicity, household income quartile, primary payer, smoking, and alcohol.

**Table 3.** Unadjusted and adjusted associations of HMB with CVD events including diabetes among hospitalized women of ages between 12-40 years; NIS-2017.

|  | Unadjusted model |  | Adjusted model* |  |
| --- | --- | --- | --- | --- |
|  | OR (95% CI) | p-value | OR (95% CI) | p-value |
| **MACE** |  |  |  |  |
| No HMB (reference) | 1 |  | 1 |  |
| Only HMB | 2.13 (1.82; 2.50) | <0.001 | 1.69 (1.43, 2.00) | <0.001 |
| HMB with IM | 1.62 (1.16; 2.27) | 0.005 | 1.28 (0.91, 1.80) | 0.158 |
| **CHD** |  |  |  |  |
| No HMB (reference) | 1 |  | 1 |  |
| Only HMB | 2.15 (1.71; 2.70) | <0.001 | 1.82 (1.43, 2.31) | <0.001 |
| HMB with IM | 2.54 (1.69; 3.81) | <0.001 | 2.10 (1.39, 3.18) | <0.001 |
| **Stroke /CVA** |  |  |  |  |
| No HMB (reference) | 1 |  | 1 |  |
| Only HMB | 1.79 (1.27; 2.53) | 0.001 | 1.53 (1.08, 2.17) | 0.017 |
| HMB with IM | 1.61 (0.76; 3.38) | 0.212 | 1.41 (0.67, 2.98) | 0.367 |
| **HF** |  |  |  |  |
| No HMB (reference) | 1 |  | 1 |  |
| Only HMB | 2.16 (1.79; 2.61) | <0.001 | 1.68 (1.38, 2.05) | <0.001 |
| HMB with IM | 1.66 (1.12; 2.47) | 0.012 | 1.27 (0.86, 1.89) | 0.232 |
| **AF/ Arrhythmia** |  |  |  |  |
| No HMB (reference) | 1 |  | 1 |  |
| Only HMB | 2.07 (1.70; 2.54) | <0.001 | 1.94 (1.58, 2.37) | <0.001 |
| HMB with IM | 2.04 (1.39; 2.97) | <0.001 | 1.89 (1.29, 2.77) | 0.001 |
| **MI** |  |  |  |  |
| No HMB (reference) | 1 |  | 1 |  |
| Only HMB | 2.12 (1.42; 3.16) | <0.001 | 1.80 (1.20, 2.69) | 0.004 |
| HMB with IM | 1.45 (0.54; 3.86) | 0.457 | 1.23 (0.46, 3.30) | 0.680 |
| **DM** |  |  |  |  |
| No HMB (reference) | 1 |  | 1 |  |
| Only HMB | 1.74 (1.59; 1.91) | <0.001 | 1.49 (1.36, 1.64) | <0.001 |
| HMB with IM | 1.49 (1.23; 1.80) | <0.001 | 1.26 (1.03, 1.54) | 0.024 |

Abbreviations: NIS, National inpatient sample; OR, Odds ratio; RR, Relative risk; CI, Confidence interval; MACE, Major adverse cardiovascular event; CHD, coronary heart disease; CVA, Cerebrovascular accident; HF, Heart failure; AF, Arterial fibrillation; MI, Myocardial infarction; DM, Diabetes; HMB, Heavy menstrual bleeding; IM, Irregular menstruation.
MACE was defined as the composite of myocardial infarction, stroke, and heart failure.
\* Adjusted model included race/ethnicity, household income quartile, primary payer, smoking, and alcohol.

### Adjusted associations of HMB diagnosis with CVD outcomes in sensitivity analyses

The adjusted association of HMB with CVD outcomes remained unchanged after additionally adjusting for obesity, MS, or DM (Table S3). These adjusted associations remained unchanged even after excluding hospitalizations with PCOS, congenital heart disease, or anticoagulation use records from the analysis (Table S4). Although age was a modifier, almost all CVD outcomes including MACE (OR=1.24, 95%CI: 1.07, 1.45, p=0.005), CHD (OR=1.33, 95%CI: 1.08,1.64, p=0.008), HF (OR=1.24, 95%CI: 1.04, 1.49, p=0.019), AF/arrhythmia (OR=1.96, 95%CI: 1.42,-2.02, p<0.001), and DM (OR=1.20, 95%CI: 1.10, 1.31, p<0.001) were significantly associated with HMB even after additionally adjusting for age, with race/ethnicity, household income quartile, primary payer, alcohol intake, and smoking status (Table S5).

## DISCUSSION

We observed a notable increase in MACE, CHD, HF, AF/arrhythmia, MI and DM associated with HMB among hospitalizations of young women. We also observed a strong age-dependent association of HMB with stroke in adjusted analyses. HMB was more likely to be associated with CVD outcomes with decreasing reproductive ages. These associations were more pronounced with HMB without IM compared to HMB with IM in hospitalized women. Unlike our study, no study directly evaluates the association of HMB with any CVD outcomes. However, one study found that women diagnosed with menorrhagia and who underwent hysterectomy had increased levels of serum inflammatory markers, predisposing them to future cardiovascular events.^25^ In another cohort study^26^, a higher risk of CVD and adverse metabolic health was observed among women who underwent a hysterectomy with ovarian conservation compared to those who did not. In addition, several studies reported that obesity and inflammation are strongly associated with HMB which may increase the risk of CVD and other associated adverse health outcomes.^10, 19, 20^ A few studies recognized that HMB is associated with IM, hirsutism, PCOS, and secondary amenorrhea among teenagers and young women which lead to insulin resistance and other metabolic abnormalities and subsequently CVD.^18, 23, 27^ In conjunction with our findings, these all studies confirm a potential risk of CVD among HMB patients.

Our study estimated an almost 1.5-fold increased prevalence of CVD outcomes associated with HMB diagnosis if adjusted analysis includes obesity and other considered factors. Although the associations between HMB and CVD outcomes were significant, the effect sizes were attenuated after additionally adjusting for age owing to a strong interaction with age. It has been observed that HMB is more prevalent in extreme age groups in reproductive ages.^10, 28^ Further analyses suggest that a higher proportion of HMB associated with CVD events occurred in hospitalized women of younger ages indicating that HMB is an early sign and critical factor for CVD in young women. However, a longitudinal study is required to confirm the age at onset of the HMB effect on the risk of CVD development. It is often considered that the co-occurrence of IM and PCOS with HMB might be a potential reason for the development of CVD outcomes. Multiple studies report the associations of IM and PCOS with CVD outcomes.^2, 15, 27^ However, HMB alone was strongly associated with most CVD outcomes while HMB with IM was profoundly associated with CHD in our study. We also observed an association between HMB and CVD outcomes after excluding PCOS cases. This finding suggests that HMB without comorbid IM or PCOS itself is an important factor for CVD outcomes. It has been shown that HMB could be a side effect of anticoagulation use.^9^ However, our analysis infers that HMB is associated with CVD outcomes regardless of anticoagulation use.

Similar to our study, advancing reproductive age, black race, lower household income, and obesity were also found to be associated with HMB in previous studies. In addition to these factors, vitamin D deficiency, dietary, environmental, and lifestyle factors, cesarean section, drug use, and parity have been found to be associated with HMB.^10, 29^ Recent studies also highlighted the impact of coronavirus disease-19 on HMB condition due to stress-induced effects on hypothalamic function as well as being presence of the virus receptor and angiotensin-converting enzyme in the women’s uterus and ovary.^30, 31^ This postulates a link between inflammation, mental health, and CVD outcomes among women with menstrual disorders, especially HMB. Although the precise mechanisms for HMB-associated cardiac abnormalities are unknown, one of the main reasons is the hormonal dysregulation related to progesterone, estradiol, glucocorticoids, and androgens leading to hypoxia, inflammation, impaired vasoconstriction, and altered hemostasis.^10^ In HMB patients, reduced expression of hypoxia-inducible factor (HIF)α, the proliferation of vascular smooth muscle and transforming growth factor β1 are linked with hypoxia and menstrual repair.^32^ Increased expression of 11β-hydroxysteroid dehydrogenase in HMB patients seem to affect cortisol and vasoconstriction.^33^ In addition, reduced expression of HOXA10 and 11 genes, altered function of estrogen receptor β and progesterone receptor, and increased matrix metalloproteinases and nuclear factor kappa B activities have been reported to influence inflammation.^34, 35^ In HMB patients, increased levels of tissue plasminogen activator affecting hemostasis, fibrin, and D-dimers have also been reported.^10, 36^ The related treatments such as hormonal-based treatments including combined oral contraceptives or progestin-only contraceptives, and hemostatic therapy affecting these pathways have been developed for HMB.^8, 9, 20^ Although the use of these treatments for HMB including hysterectomy depends on multiple factors age, HMB pathology, fertility status, and desire to become pregnant^9^, shared decision making and effective treatment strategies are required for CVD management associated with HMB.^8, 10, 37^ In addition, reduction of environmental exposure is also useful for minimizing risk of menstrual disorders and CVD events.^3, 38^ A higher prevalence of HMB has been reported in women with congenital cardiovascular events. ^39, 40^ In our study, the association remained consistent between HMB and CVD outcomes even after excluding congenital cardiovascular events. Our sensitivity analysis also indicates that the association of HMB with CVD outcomes was independent of obesity, metabolic syndrome, DM, and PCOS. This suggests that HMB may be associated with CVD outcomes through stress-induced inflammation, oxidative stress and inflammation, iron deficiency, iron deficiency anemia, and hemodynamic instability.

Our study is a cross-sectional study design using hospitalization records which is a major limitation since it limits the generalizability of our study findings. Due to retrospective analysis of a secondary database, we were unable to account for some confounders including type of anticoagulants and tranexamic acid use, dietary pattern, oral contraceptive use, family history, or other comorbidities including the onset of menorrhagia. Moreover, ICD-10 codes may not fully capture MS and may underestimate HMB diagnosis among all hospitalizations leading to biased associations. Despite these limitations, our study attempted to address a critical question for improving women’s CVD health utilizing a large population database representing US hospitalizations. The comprehensive inclusion of CVD events and rigorous analyses including stratified analysis of HMB with or without IM with multiple sensitivity analyses based on obesity, MS, DM, PCOS, and congenital heart disease provide valuable contributions to the limited literature on HMB and CVD health. Furthermore, the moderate to large effect sizes for the association between HMB and CVD events obtained in our study signify the early screening and management of HMB for the prevention of CVD events.

## CONCLUSIONS

In our study of US hospitalizations of young women, HMB was found to be strongly associated with CVD events independent of obesity, MS, and DM. HMB without IM was profoundly associated with most CVD outcomes. Although younger ages of HMB seem to be an early indication for adverse CVD outcomes, this requires further investigation. Future cohort studies are required to evaluate the long-term effect of HMB on the development of CVD outcomes. Our findings suggest promoting increased awareness, early detection, and optimum management of HMB to avoid adverse consequences of HMB including CVD outcomes among pubertal and reproductive-aged females.

## Data Availability

All the study data are available at National (Nationwide) Inpatient Sample (NIS) and can be purchased and access through the website: https://hcup-us.ahrq.gov/nisoverview.jsp

## Acknowledgments

The authors acknowledge the Office of Research resources for providing suitable assistance with the statistical work.

## Sources of Funding

None

## Disclosures

All authors have nothing to disclose.

## Supplemental Material

Tables S1-S5

## Notes

### Competing Interest Statement

The authors have declared no competing interest.

### Clinical Trial

Not applicable.

### Funding Statement

No external funding was received for this study.

### Author Declarations

The NIS, 2017 includes de-identified datasets from the International Classification of Diseases, 10th Revision (ICD-10) codes from participating hospitals and hence, does not require participant consent or approval.

